# Privacy-Preserving, Decentralised Inference of Epidemic Transmission Dynamics from Digital Exposure Histories

**DOI:** 10.64898/2026.10.05.26364757

**Authors:** Younjung Kim, Ben Lambert, Luca Ferretti, Michelle Kendall, Robert Hinch, Jamie Webb, Thomas Rawson, Lucie Abeler-Dörner, Peter Horby, Michael Parker, Melinda C. Mills, Christophe Fraser, Moritz U.G. Kraemer, Christl A. Donnelly

## Abstract

**Background:** Exposure-history data collected during epidemics or pandemics within digital contact-tracing applications hold immense potential for rapid, scalable epidemiological inference. The sensitivity of these data makes privacy-preserving designs essential to secure public trust and app uptake. However, during the COVID-19 pandemic, such approaches were not yet available at the scale or maturity required for deployment. We present a Bayesian decentralised framework to infer transmission dynamics from exposure-history data while protecting user privacy.

**Methods:** Our approach is based on iterative communication between user devices and a central server, computing log-likelihood gradients on device from exposure-history data and centrally updating parameter values using only aggregated gradients. Applying this framework to synthetic exposure-history data from a hypothetical respiratory pathogen epidemic simulated on dynamic contact networks, we assessed its ability to estimate the underlying transmission parameters, perform infector attribution, and reconstruct serial-interval (SI) distributions. We systematically explored how inference was affected by incomplete exposure-history data arising from partial app adoption, case under-ascertainment, and incomplete exposure logging, and by transmission scenarios.

**Findings:** Our decentralised framework accurately recovered transmission parameters, closely matching estimates derived from centralised data with no systematic bias in posterior means. Parameter recovery remained robust when hazards from unobserved exposures were known as a daily average across the population. Although incomplete exposure-history data and specific transmission scenarios introduced compensatory parameter biases, the inferred host infectiousness profile retained sufficient accuracy, and the weekly SI distributions reconstructed from infector attributions closely resembled the ground-truth distributions.

**Interpretation:** Our findings highlight that, beyond its primary role as a control measure, digital contact tracing could help characterise transmission dynamics almost in real time and inform public health interventions while preserving user privacy.

**Funding:** Oxford Martin School Programme on Digital Pandemic Preparedness; National Institute for Health and Care Research (NIHR) Health Protection Research Unit in Emerging and Zoonotic Infections.

**Evidence before this study:** We searched PubMed for studies published from database inception to Sept 30, 2026, using the keywords (“digital contact tracing” OR “exposure notification” OR “contact tracing app”) AND (“federated learning” OR “decentralised” OR “privacy-preserving” OR “statistical inference” OR “Bayesian inference”) AND (“transmission” OR “serial interval” OR “infectiousness”). During the COVID-19 pandemic, digital contact-tracing (DCT) apps were deployed at an unprecedented scale: 224 COVID-19 apps were launched across 127 countries, 128 of which supported exposure notification. In England and Wales, the NHS COVID-19 app had 16.5 million regular users (28% of the total population) and averted an estimated 1 million cases and 9,600 deaths in its first year. To protect privacy, however, most apps used decentralised architectures that kept exposure-history data on users’ devices. This privacy-preserving principle has largely prevented these data from being used for epidemiological inference to understand transmission dynamics, such as estimating the serial-interval (SI) distribution and host infectiousness profile over time. While existing decentralised algorithms estimate each user’s infection risk, we found no study that developed and evaluated a framework for estimating population-level epidemiological parameters from exposure-history data that remain on users’ devices.

**Added value of this study:** To our knowledge, this is the first such framework. It estimates key quantities required for understanding transmission dynamics, including how an infected individual’s infectiousness changes over time relative to their symptom onset date, how much close contacts increase the risk of transmission, and the SI distribution. The framework learns an exposure hazard function from data that never leave users’ devices, then attributes infections to likely infectors, and reconstructs the SI distribution. We applied the framework to synthetic exposure-history data from epidemics simulated on dynamic contact networks of 166,034 individuals, matching the 2024 population of Oxford, UK. It produced estimates comparable to those from centralised analysis, identified the timing of peak host infectiousness relative to symptom onset to within one day, and reconstructed SI distributions that closely tracked the ground truth. We also systematically assessed how participation levels (app adoption and case ascertainment), technical limitations (incomplete exposure logging), and surveillance (knowledge of the background hazard from unobserved exposures) affected inference, and showed that daily population-level knowledge of the background hazard was sufficient for reliable inference.

**Implications of all the available evidence:** Beyond their role as a control measure, DCT apps could provide timely, population-representative estimates of key transmission parameters, such as the host infectiousness profile and SI distribution. Our findings show that such inference is feasible within the privacy-preserving architectures likely to underpin future DCT systems. Greater app adoption, case ascertainment, and exposure logging would improve inference accuracy. To realise this potential, the framework will need to be embedded in DCT systems, supported by concurrent epidemiological research, real-world validation, and early collaboration with public health authorities and platform providers.

## 1. Introduction

During the COVID-19 pandemic, digital contact-tracing (DCT) applications were deployed globally at an unprecedented scale^1,2^. DCT apps detect potentially infectious contacts (exposures) by exchanging rolling proximity identifiers with nearby devices via Bluetooth, enabling rapid and scalable notification, isolation, and testing of exposed individuals^3,4^. The resulting digital exposure-history data hold substantial epidemiological value, recording high-resolution proxies for key transmission determinants: (a) Bluetooth signal strength (for contact proximity), (b) signal exchange time (for contact duration), and (c) the time between exposure and the contact’s symptom onset, if reported.

These data could prove particularly valuable for inferring the serial-interval (SI) distribution (an SI is the time interval between symptom onsets within a transmission pair). The SI distribution is critical for characterising transmission dynamics, including estimating reproduction numbers assuming it approximates the generation-time (GT) distribution (a GT is the time interval between infections within a transmission pair)^5^. Although the SI distribution can vary during an epidemic^6–8^, it is typically estimated in small, context-specific settings^8,9^. Moreover, its estimation often relies on individuals with clear links to their infectors, whose exposure patterns may not be representative of those of the general population^10^. Consequently, estimated distributions may not reflect population-wide dynamics, potentially biasing downstream estimates^8^. In contrast, digital exposure-history data could provide more timely and representative estimates if analysed within frameworks that link each infection to its true infector or to a potential infector whose exposure occurred at a similar point in their own infectiousness profile to that of the true infector.

Another potential application is elucidating intrinsic host infectiousness profiles. Studies of within-host pathogen dynamics can track individuals’ pathogen load over time but are limited by small sample sizes, uncertain infection times, and the imperfect correlation between pathogen load and host infectiousness^11,12^. In contrast, digital exposure-history data could reveal when transmission is most likely relative to symptom onset by capturing exposures and resulting infections across each infected individual’s disease course.

Despite this potential, digital exposure-history data were heavily under-utilised for epidemiological inference during the COVID-19 pandemic, mainly because most DCT apps employed decentralised architectures that kept privacy-sensitive exposure-history data on users’ devices, a practice likely to remain standard in future implementations^1^. This highlights an urgent need for analytic frameworks that operate within decentralised architectures.

Here, we propose federated learning as a promising approach for epidemiological inference from privacy-sensitive exposure-history data. Federated learning operates through iterative communication between a central server and local devices: data are processed locally, and only aggregated, de-identified information is sent to the server to update model parameters^13^. This framework opens a new avenue for privacy-preserving epidemiological inference without centralising exposure-history data, provided that appropriate statistical signals can be derived on local devices and shared with the central server.

To assess this potential, we developed a Bayesian federated-learning framework to infer transmission dynamics at the exposure, individual, and population levels. We evaluated this framework using synthetic digital exposure-history data, generated by simulating the spread of a hypothetical respiratory pathogen on dynamic contact networks. From these data, we estimated the parameters of the exposure hazard while accounting for incomplete exposure records. Using these estimates, we then identified the most likely infector for each user reported as infected and reconstructed the SI distribution based on these attributions across the population. We evaluated the robustness of our inferences across transmission and inference scenarios varying (a) the level of the background hazard from unobserved exposures, (b) knowledge of this background hazard, and (c) the underlying transmission dynamics.

## 2. Methods

### 2.1. Epidemic Simulation for Exposure-History Data Generation

Using OpenABM-Covid19^14^, we generated dynamic daily contact networks for 166,034 simulated individuals, matching the 2024 population of Oxford, a representative mid-sized UK city. These networks, parameterised to the UK context, included age-stratified mixing across household, occupational (e.g. workplaces for working-age adults, schools for children, or regular social activities for older adults), and random (e.g. public transport, social gatherings) settings^14^, with a mean of 12.5 daily contacts per person^15,16^.

The epidemic simulation on these networks assumed a simple SIR structure with a daily time step, comprising susceptible (S), infected and infectious (I), and recovered (R) states. Without using a separate exposed state, the latent period was captured through the host infectiousness profile, defined relative to each individual’s symptom onset. All infected individuals developed symptoms and were classified as cases or non-cases according to a predefined level of case under-ascertainment. Neither symptom onset nor case status affected contact patterns, and recovered individuals retained immunity for the remainder of the simulation.

We simulated the transmission of a hypothetical respiratory pathogen over 365 days on each network using a log-linear hazard function (Table 1). For each exposure, this function combined four parameters (the baseline hazard, the proximity-effect coefficient, and the mean and standard deviation of the host infectiousness profile), with exposure-specific inputs: proximity and duration, and the potential infector’s symptom onset date, sampled from assumed or empirical distributions (Table 1).

**Table 1.** Epidemic simulation parameters, units, and input data, based on the hazard function: *h_e_*(*τ*) = exp (*β*_baseline_ + *β*_proximity_ · ρ_e_) · *ω*(τ — *o_i_*) · *d_e_* (building upon the formulae used to compute transmission scores by the National Health Service [NHS] COVID-19 app^30^)

| Parameter / data | Description | Unit | Transmission scenarios |  |
| --- | --- | --- | --- | --- |
|  |  |  | Baseline | Alternative |
| Primary parameters |  |  |  |  |
| $R_0$ | The expected number of secondary cases produced by a single infected individual in a completely susceptible population. With $\beta_{\text{proximity}}$ and $\omega(\cdot)$ determined, $\beta_{\text{baseline}}$ was calibrated to achieve $R_0$. | Dimensionless | 1.25 | 2.5, 5.0 |
| $\gamma_{\text{proximity}}$ | Sets $\beta_{\text{proximity}}$ to scale hazard per one-standard-deviation increase in proximity | Dimensionless | 2 | 1, 4 |
| $\mu_{\text{infectiousness}}$ | Shifts peak of host infectiousness profile $\omega(\cdot)$ relative to symptom onset | Days | 0 | -2, 2 |
| $\sigma_{\text{infectiousness}}$ | Controls temporal dispersion of host infectiousness profile $\omega(\cdot)$ | Days | 2 | 1, 4 |
| $\mu_{\text{incubation}}$ | Shifts mean incubation period; variance held constant | Days | 6.4 | 3.2, 12.7 |
| $\delta$ | Introduces moderate or strong overdispersion in host infectiousness | Dimensionless | 0 | 0.35, 0.70 |
| Derived parameters and functions |  |  |  |  |
| $\beta_{\text{baseline}}$ | Baseline log-hazard per day of contact. Calibrated to achieve target $R_0$ | $\log(\text{day}^{-1})$ | Derived from $R_0$ | |
| $\beta_{\text{proximity}}$ | Log-hazard ratio per unit increase in $\rho_e$ | Dimensionless | Derived from $\gamma_{\text{proximity}}$ | |
| $\omega(\cdot)$ | Host infectiousness profile defined by a peak-normalised normal distribution, centred such that 0 marks symptom onset (mean: $\mu_{\text{infectiousness}}$, standard deviation: $\sigma_{\text{infectiousness}}$) | Dimensionless (0–1) | Derived from $\mu_{\text{infectiousness}}$ & $\sigma_{\text{infectiousness}}$ | |
| Input data |  |  |  |  |
| $\rho_e$ | Proximity of exposure $e$. Sampled from a beta distribution to reflect contact patterns in which non-physical, casual contacts form the majority of interactions<sup>31</sup>. | Dimensionless (0–1) | Beta (2, 5) | |
| $d_e$ | Duration of exposure $e$. Sampled from a radio frequency identification (RFID)-based co-location empirical dataset<sup>32</sup>. Each pair's cumulative daily contact duration was rounded up to the nearest multiple of 30 minutes and expressed as a number of 30-minute windows, mirroring the maximum length of the exposure windows used by the NHS COVID-19 app to assess transmission risk<sup>30</sup>. $d_e$ proportionally scales hazard. | Fraction of a day (in 30-min increments) | Génois and Barrat (2018)<sup>32</sup> | |
| $o_i$ | Infectors $i$'s symptom onset date. Sampled from an empirical incubation-period distribution<sup>18</sup>. | Calendar day | Gamma (2.29, 0.36), $\mu_{\text{incubation}}=6.4^{18}$ | Gamma (2.29, 0.72), $\mu_{\text{incubation}}=3.2$; Gamma (2.29, 0.18), $\mu_{\text{incubation}}=12.7$ |

Each day, transmission to each susceptible individual was determined by a Bernoulli trial based on their total daily hazard. If infection occurred, the true infector was sampled from the contributing exposures with probabilities proportional to their hazards. For each newly infected individual, a symptom onset date was drawn from an assumed incubation-period distribution, which then determined their intrinsic infectiousness on subsequent days (Supplementary Information pp 2–3).

Epidemics were simulated under various ‘transmission scenarios’ (Table 1). Under the baseline transmission scenario, parameters for intrinsic host infectiousness and the incubation-period distribution mirrored those of the SARS-CoV-2 virus, the causative agent of COVID-19^3,17,18^. The proximity-effect coefficient was set so that a one-standard-deviation increase in the proximity score doubled the hazard. The baseline hazard was calibrated to yield a basic reproduction number (*R*_0_) of 1.25, reflecting a regime where targeted interventions such as DCT remain viable and are the primary means of control^18^. We also considered alternative transmission scenarios to assess their impact on inference (Table 1). For each scenario, we ran 100 independent simulations, each generating one exposure-history dataset.

### 2.2. Decentralised Epidemiological Inference Framework

We estimated the parameters of the underlying hazard function by applying a Bayesian federated-learning framework to the simulated exposure-history data (Fig. 1) and evaluated its robustness under plausible data-availability scenarios (hereafter ‘inference scenarios’; Table 2).

**Figure 1.**
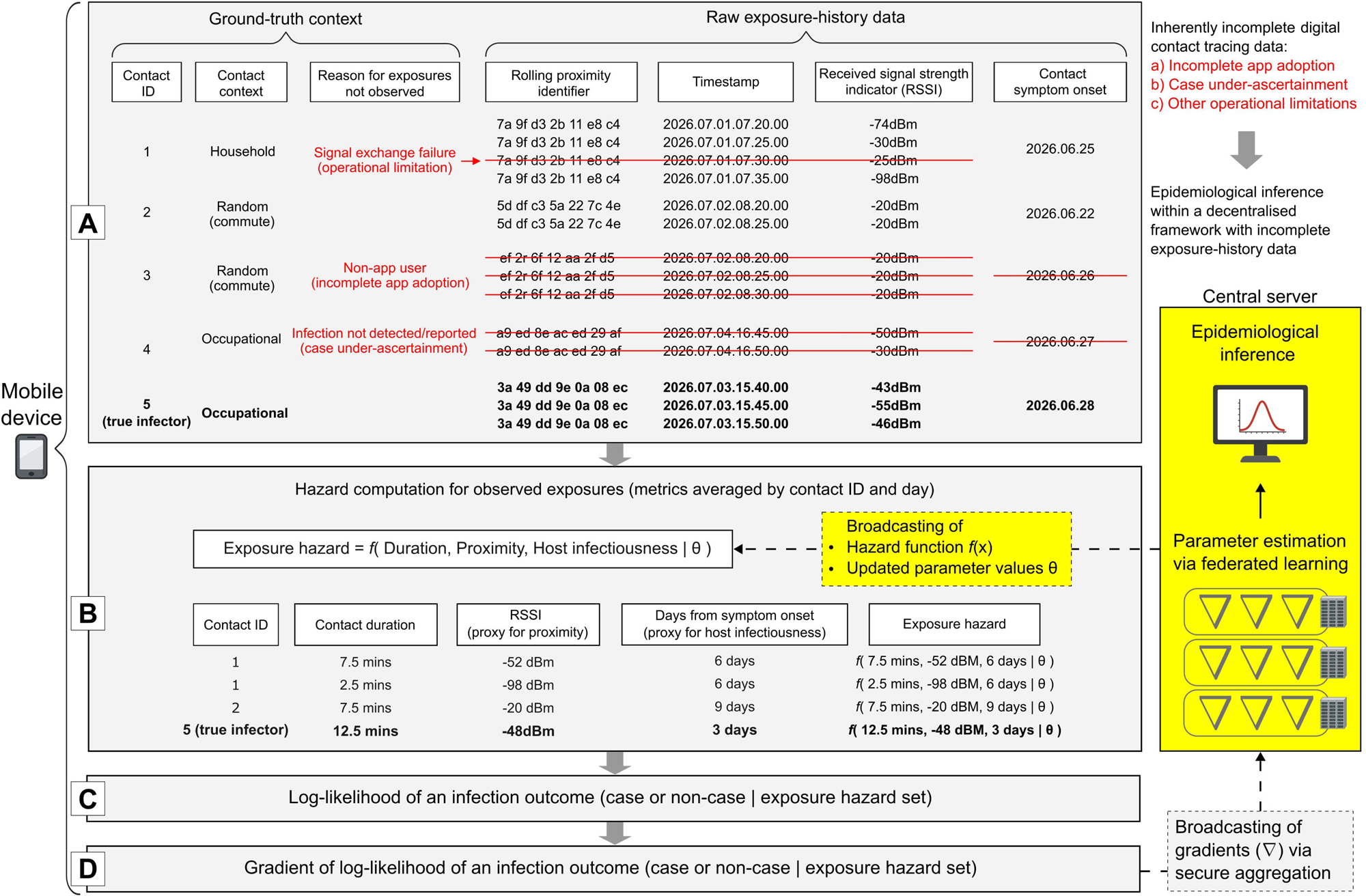
Decentralised analytic framework for epidemiological inference from digital exposure-history data. Processes are partitioned between local mobile devices (grey background; Boxes A–D) and a central server (yellow background), which estimates transmission parameters via federated learning and conducts downstream inferences. Dotted boxes show data communicated to and from the central server, coloured by source (yellow, server; grey, mobile device). (A) Ground-truth context of potentially infectious contacts (exposures) and the corresponding raw exposure-history data, logged via Bluetooth signal exchange between mobile devices. Red strikethroughs mark exposures that go unobserved due to inherently incomplete contact tracing, specifically: (a) incomplete app adoption, (b) case under-ascertainment, or (c) other operational limitations. Bold text indicates the true infector. (B) Exposure hazard for each observed exposure, computed from contact duration, received signal strength (a proxy for proximity), and days since the contact’s symptom onset (a proxy for intrinsic host infectiousness), using the hazard function and parameter values broadcast by the central server. (C) Log-likelihood of the device user’s case status (case or non-case) given the set of exposure hazards, and (D) its gradient. The central server receives these gradients via secure aggregation, iteratively updates the parameter values and broadcasts them back to mobile devices, and uses the final estimates for downstream inference. Data shown are illustrative.

**Table 2.** Epidemiological inference from simulated epidemic datasets.

| Category | Description | Inference scenarios |  |
| --- | --- | --- | --- |
|  |  | Baseline | Alternative |
| a. Completeness of digital exposure-history data |  |  |  |
| App adoption | Percentage of the population using a DCT app and sharing their digital exposure-history data | 20% | 40–100% |
| Case ascertainment | Percentage of infected individuals identified as cases by the DCT app | 100% | 25–75% |
| Technical limitation | Percentage of exposures that are not logged due to technical limitations or user usage pattern | 0% | 25–75% |
| b. Knowledge about background hazard from unobserved exposures |  |  |  |
| Background hazard knowledge | Scenario 1: Perfect individual- and daily-level background hazard knowledge<br>Scenario 2: Daily population-averaged background hazard knowledge<br>Scenario 3: Uniform background hazard across individuals and time | Scenario 2 | Scenarios 1 & 3 |

First, exposure-history data are typically incomplete, primarily due to partial app adoption, case under-ascertainment, and operational limitations affecting exposure logging (e.g. Bluetooth deactivation or devices being switched off). Under the baseline inference scenario, we randomly designated 20% of individuals as app users and retained only exposures between two app users (comparable to the 23% adoption observed in England and Wales shortly after the launch of the NHS COVID-19 app^4^). We also explored higher adoption levels. The baseline scenario assumed complete case ascertainment and exposure logging in all instances when one app user exposured another app user; alternative scenarios relaxed these assumptions with app adoption held at 20% (Table 2).

Second, the incompleteness of app adoption, case ascertainment, and explore logging leaves some exposures unobserved. Knowledge of the resulting ‘background hazard’ can vary with surveillance capacity and understanding of app usage patterns. We tested three scenarios (Table 2, Supplementary Information pp 3–4). Scenario 1 assumes an idealised situation, where background hazards are known at both the daily and individual levels. In Scenario 2, the background hazard is known only at the daily population level. We selected Scenario 2 for our baseline inference scenario because the required information could be approximated from population-level surveillance data (e.g. prevalence and incidence). Finally, Scenario 3 represents the least-informed situation, in which no information on epidemic dynamics is available, so the background hazard is assumed constant throughout the epidemic.

Third, exposure-history data must remain on local devices. Because standard Markov chain Monte Carlo (MCMC) approaches require full-data likelihood evaluations, they are infeasible in a decentralised setting. We therefore adopted a federated-learning framework using stochastic variational inference (SVI)^19^ to optimise the parameters of a global multivariate normal approximation to the posterior (i.e. its mean vector and covariance matrix), capturing both parameter uncertainties and correlations (Supplementary Information pp 4–7). In each iteration, acting as the central server, we sampled parameter values from the global distribution and conceptually broadcast them to a random mini-batch of app users. Then, acting as local devices, we computed the log-likelihood gradient for each sampled user given their case status (case or non-case), exposure-history data, the pre-shared hazard function and background-hazard data, and the sampled parameter values. Acting again as the central server, we aggregated these local gradients to estimate the gradient of the evidence lower bound (ELBO) and updated the global variational parameters using the Adam optimiser^20^. This cycle was run for 5000 iterations, and final estimates were obtained by averaging the variational means over the last 10% of iterations. For the baseline transmission and inference scenarios, we also applied a secure aggregation process, masking individual users’ raw gradients, to assess whether individual privacy (e.g. case status and exposure patterns) was protected without compromising inference^21^ (Supplementary Information pp 7–8).

The log-likelihood functions incorporated the same hazard function as the epidemic simulation for observed exposures, with an additional term representing the background hazard from unobserved exposures (Supplementary Information pp 2–4). Their form depended on whether an app user was observed as a case or a non-case on the day of analysis, *T*. A case was an infected user who developed symptoms on day *o_j_* < *T*, with their log-likelihood summing the joint probability of infection on day *τ* and an incubation period of *o_j_* — *τ* across all days up to symptom onset. A non-case was a user who showed no symptoms by day *T*, being either never infected or infected but not yet symptomatic. Their log-likelihood combined the probability of remaining uninfected by day *T* with the summed probabilities of being infected on any day up to day *T* followed by symptom onset after day *T*. Because the infection day (*τ*) was unknown, both log-likelihood functions accounted for all possible infection days, with the probability of infection on any day *τ* conditional on the user having remained uninfected on all prior days.

### 2.3. Evaluation of Epidemiological Inference

Based on the estimated parameters, we attributed infections to either observed or unobserved exposures as follows. For a given case, using the same inference model, we first determined the infection day and then attributed the infection to an exposure observed on that day or to the background hazard, with probabilities proportional to their respective hazards. We also reconstructed SI distributions from hazard-based attributions to observed exposures. We evaluated these inferences in three ways, aggregating results across the simulated datasets for each combination of transmission and inference scenarios. First, we assessed parameter recovery by comparing our estimates with the ground-truth values and with estimates obtained from centralised exposure-history data using Hamiltonian Monte Carlo (HMC) with the rstan package^22^ (Supplementary Information p 8). Second, we evaluated the accuracy of hazard-based infector attributions in terms of specificity, sensitivity, and sensitivity relative to non-hazard-based attributions. Finally, we compared SI distributions reconstructed from hazard-based attributions with those from true transmission pairs and from non-hazard-based attributions.

## 3. Results

Estimates from our decentralised framework closely matched those from centralised exposure-history data (Fig. S1). Although posterior means showed no apparent systematic bias for any parameter, the decentralised framework yielded slightly smaller posterior standard deviations, consistent with the well-documented tendency of variational inference to underestimate posterior variance^23^.

Log-likelihood gradients differed markedly by case status across all estimated parameters (Fig. S2). Gradients from cases were positive, pulling the estimated hazards upwards and increasing the likelihood of infection given their observed exposures, whereas gradients from non-cases were consistently negative, pulling the hazards downwards. These differences were obscured after secure aggregation, which masked individual gradients to reveal only their sum to the central server (Fig. S2).

Parameter recovery varied across transmission and inference scenarios. When the background hazard was known at both the daily and individual levels (Scenario 1), parameters were well recovered across all app adoption levels, with only slight underestimation of the timing of peak host infectiousness (*μ_infectiousness_*) and correspondingly minor overestimation of the baseline hazard (*β_baseline_*) (Fig. 2). These biases persisted even at full adoption under the explored transmission scenarios and increased with *R*_0_ (Fig. S3). This was likely because many exposures, particularly repeated contacts (e.g. between household members), made infection probable soon after the infector became infectious, regardless of the exact timing of peak host infectiousness, limiting the information available to identify this peak timing. In contrast, when the background hazard was known only at the daily population level (Scenario 2) or assumed constant throughout the epidemic (Scenario 3), bias increased as app adoption decreased, although Scenario 2 estimates remained substantially more robust than Scenario 3 estimates.

**Figure 2.**
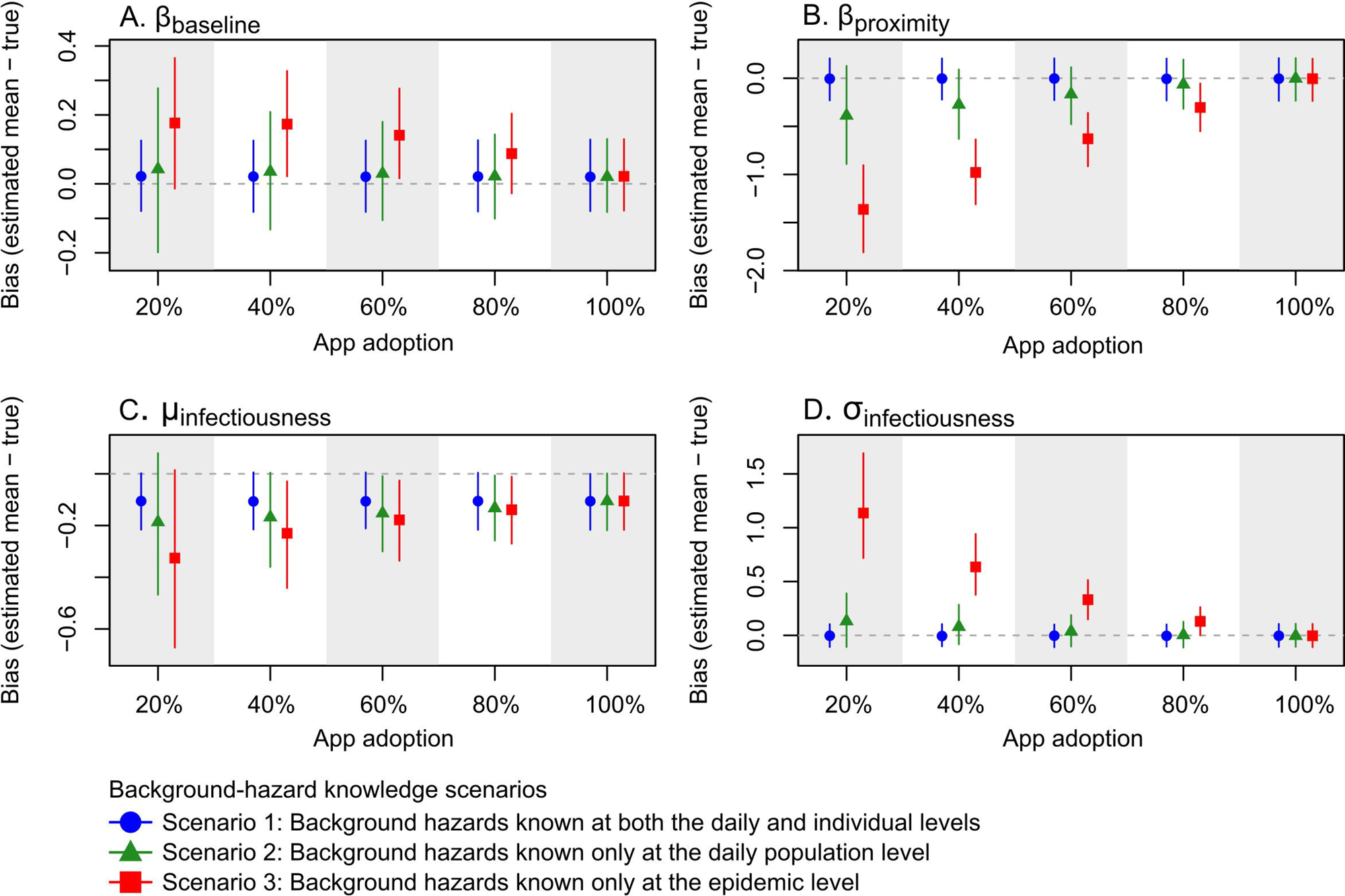
Impact of app adoption and background-hazard knowledge scenarios on parameter estimation. Bias in the estimates of four transmission parameters: (A) *β_baseline_*, the log baseline hazard per day of contact; (B) *β_proximity_*, the log hazard ratio per unit increase in the proximity score; (C) *μ_infectiousness_*, the timing of peak host infectiousness relative to symptom onset (days); and (D) *σ_infectiousness_*, the temporal dispersion of host infectiousness (days). Bias was calculated as the difference between the estimated mean and the true parameter value and is shown for each level of app adoption under three background-hazard knowledge scenarios (see legend). Points and vertical lines indicate the median and 95^th^ percentile intervals of the bias across 100 simulated exposure-history datasets. All other transmission and inference settings were held at their baseline values.

Overall, the model compensated for incomplete exposure-history data by increasing the baseline hazard (overestimated *β_baseline_*), weakening the proximity effect (underestimated *β_proximity_*), broadening the temporal dispersion of host infectiousness (overestimated *σ_infectiousness_*), and shifting peak host infectiousness earlier (underestimated *μ_infectiousness_*) (Fig. 2). These shifts were reflected in the host infectiousness profile reconstructed from the estimated *μ_infectiousness_* and *σ_infectiousness_* (Fig. 3). The profile’s peak relative to symptom onset was identified within one day in all three scenarios. Its width remained almost identical under Scenarios 1 and 2 but widened progressively as app adoption decreased under Scenario 3, reflecting the greater overestimation of *σ_infectiousness_* when background-hazard knowledge was limited (Fig. 3).

**Figure 3.**
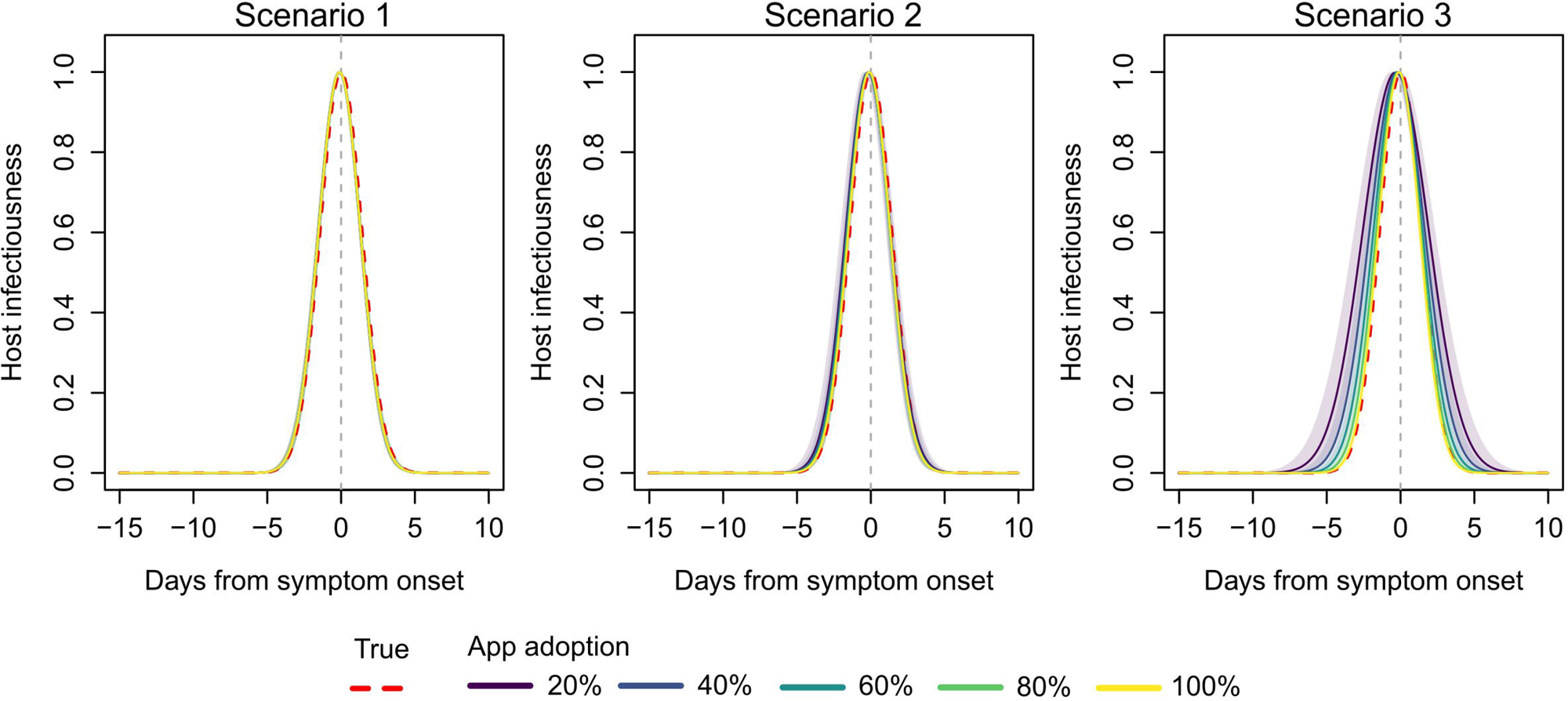
Impact of app adoption and background-hazard knowledge scenarios on the inferred host infectiousness profile. Temporal host infectiousness profiles reconstructed from the estimated *μ_infectiousness_*, the timing of peak host infectiousness relative to symptom onset (days), and *σ_infectiousness_*, the temporal dispersion of host infectiousness (days), under five levels of app adoption and three background-hazard knowledge scenarios: background hazards known at both the daily and individual levels (Scenario 1), only at the daily population level (Scenario 2), or only at the epidemic level (Scenario 3). Host infectiousness is normalised to its peak and plotted against days from symptom onset (grey dashed vertical line). Red dashed lines denote the true temporal host infectiousness profile used to simulate the epidemic. Solid lines indicate the median inferred profile across 100 simulated exposure-history datasets, coloured by the level of app adoption, and shaded regions show the corresponding 95^th^ percentile intervals. All other transmission and inference settings were held at their baseline values.

Beyond app adoption, parameter bias increased with case under-ascertainment or incomplete exposure logging (Fig. 4). Case under-ascertainment mainly caused underestimation of the baseline hazard *β_baseline_* to reconcile the exposure-history data of true cases misclassified as non-cases (Fig. 4; blue circles), whereas incomplete exposure logging biased parameters in the same direction as incomplete app adoption (Fig. 4; red triangles). Estimates remained generally robust across alternative transmission scenarios, although certain extreme values of *R*_0_, *σ_infectiousness_*, and *γ_proximity_* introduced modest biases (Fig. S3).

**Figure 4.**
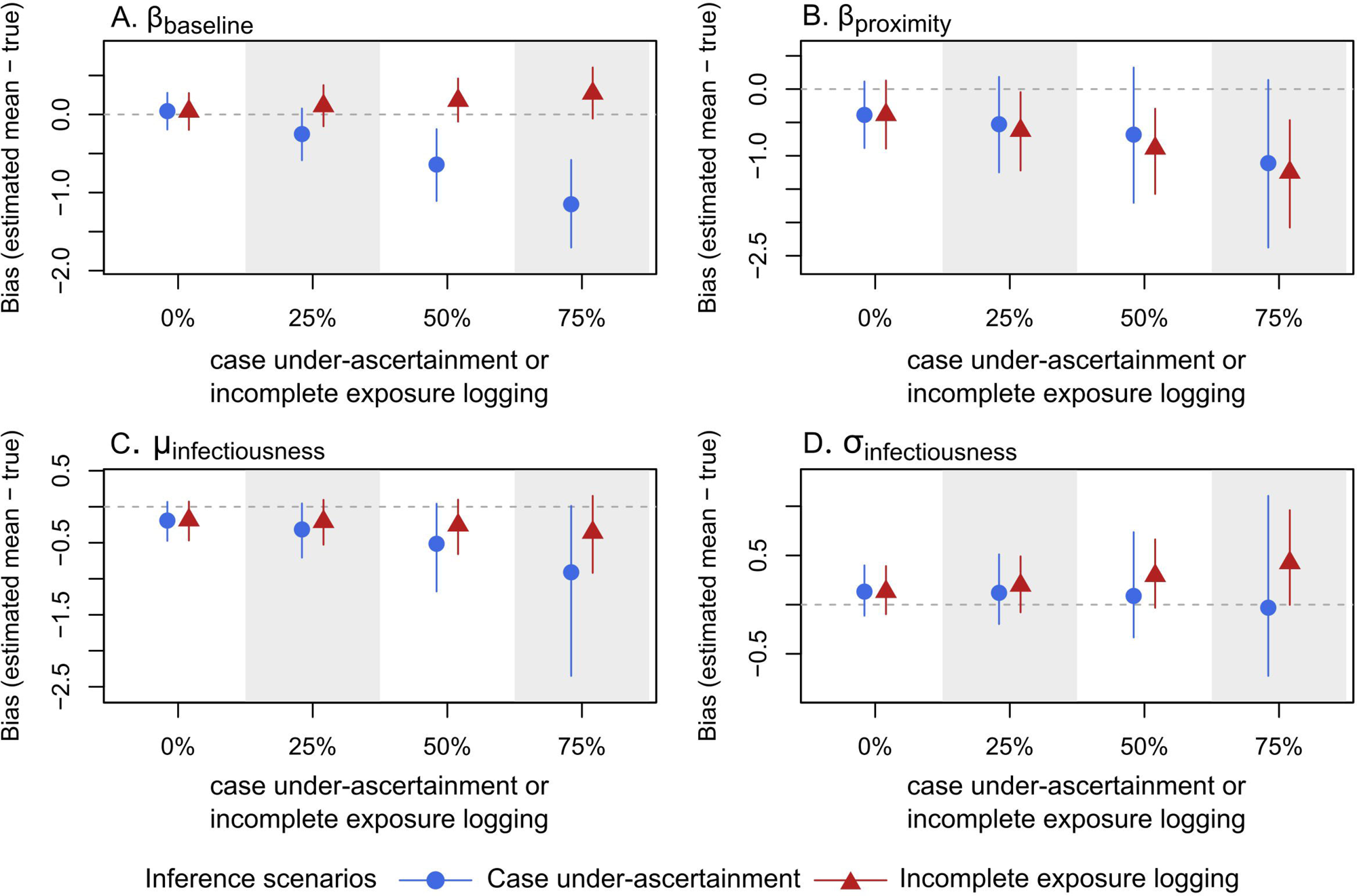
Impact of case under-ascertainment and incomplete exposure logging on parameter estimation. Bias the estimates of four transmission parameters: (A) *β_baseline_*, the log baseline hazard per day of contact; (B) *β_proximity_*, the log hazard ratio per unit increase in the proximity score; (C) *μ_infectiousness_*, the timing of peak host infectiousness relative to symptom onset (days); and (D) *σ_infectiousness_*, the temporal dispersion of host infectiousness (days). Bias, calculated as the difference between the estimated mean and the true parameter value, is shown for increasing levels of case under-ascertainment (blue) or incomplete exposure logging (red). Points show the median bias across 100 simulated exposure-history datasets, and vertical lines show the 95^th^ percentile intervals. All other transmission and inference settings were held at their baseline values, with 20% app adoption and background hazards known at the daily level (Scenario 2).

Attribution performance also varied across scenarios. Under the baseline scenario, attribution specificity (the proportion correctly attributed to the background hazard) and sensitivity (the proportion correctly attributed to the true infector among attributions made to observed exposures) were high at 0.83 (95^th^ percentile interval [PI]: 0.78–0.88) and 0.88 (95PI: 0.86–0.89), respectively (Fig. 5). Hazard-based attribution identified true infectors 1.84 times more frequently than non-hazard-based attribution. As app adoption increased, this advantage grew further, despite both attribution specificity and sensitivity decreasing owing to the growing volume of observed (and therefore competing) exposures (Fig. 5). As background-hazard knowledge decreased from Scenario 1 to Scenario 3, specificity and sensitivity both decreased (likely due to parameter biases), whereas the number of observed potential infectors increased (likely driven by a compensatory widening of the estimated host infectiousness profile) (Fig. 5). The alternative transmission scenarios that most affected parameter recovery also affected attribution. For example, an elevated *R*_0_ decreased both specificity and sensitivity but substantially expanded the pool of observed potential infectors. This expansion reduced the likelihood of correct non-hazard-based attribution more sharply than hazard-based attribution sensitivity, thereby increasing the relative sensitivity of hazard-based attribution (Fig. S4).

**Figure 5.**
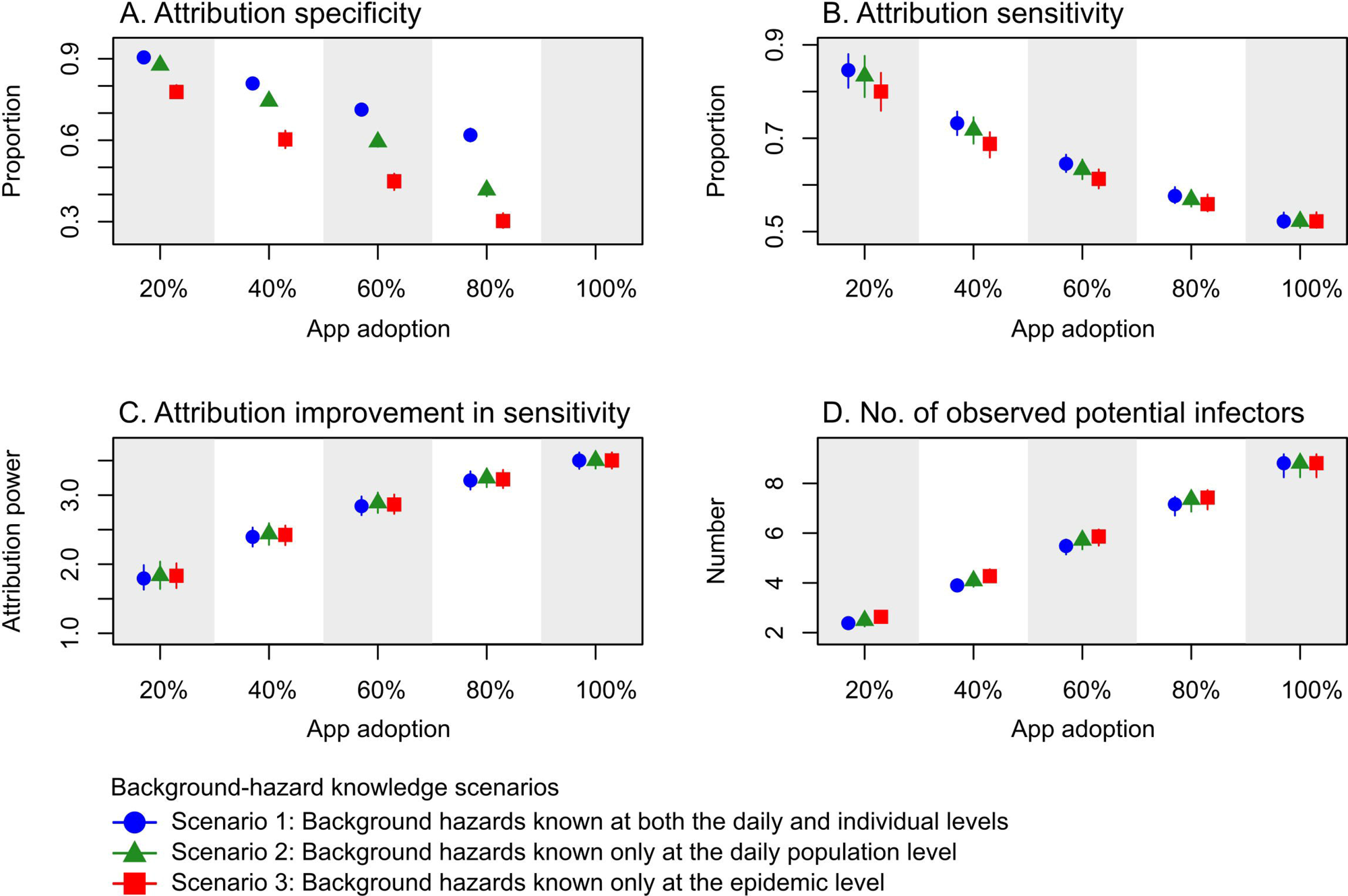
Impact of app adoption and background-hazard knowledge scenarios on infector attribution. This figure illustrates the performance of infector attribution across varying levels of app adoption and background-hazard knowledge. The four panels show: (A) attribution specificity (attribution to the background hazard when the true infector is unobserved); (B) attribution sensitivity (attribution to the true infector when attribution is made to observed exposures); (C) attribution improvement in sensitivity (improvement in sensitivity for hazard-based attribution relative to non-hazard-based attribution); and (D) the number of observed potential infectors (see the legend for definitions). Points and vertical lines represent the median and 95^th^ percentile intervals of these metrics across 100 simulated exposure-history datasets under Scenario 1 (blue, background hazards known at both the daily and individual levels), Scenario 2 (green, background hazards known only at the daily population level), and Scenario 3 (red, background hazards known only at the epidemic level). Inference was conducted under the baseline transmission and inference scenarios, with the only exceptions being the specific app adoption level and a background-hazard knowledge scenario evaluated.

Under the baseline scenario, the SI distribution reconstructed from hazard-based attribution closely matched the ground-truth SI distribution, assessed using backward SIs for transmission pairs in which infections were attributed to observed exposures (Fig. 6). Except early and late in the epidemic, when few transmission pairs were available, both its mean and uncertainty closely tracked those of the ground truth. In contrast, the SI distribution reconstructed from non-hazard-based attribution failed to track the ground truth, with mean biases of several days and substantially greater uncertainties around the epidemic peaks. The hazard-based SI distribution remained robust under alternative transmission scenarios (Fig. 7). Finally, the advantage of hazard-based attribution over non-hazard-based attribution for reconstructing the SI distribution narrowed as the level of case under-ascertainment or incomplete exposure logging increased, likely because fewer spurious exposures were available for attribution (Fig. S5). Nevertheless, while mean SIs from non-hazard-based attribution moved closer to the ground truth, their uncertainty remained substantially greater than that of hazard-based estimates.

**Figure 6.**
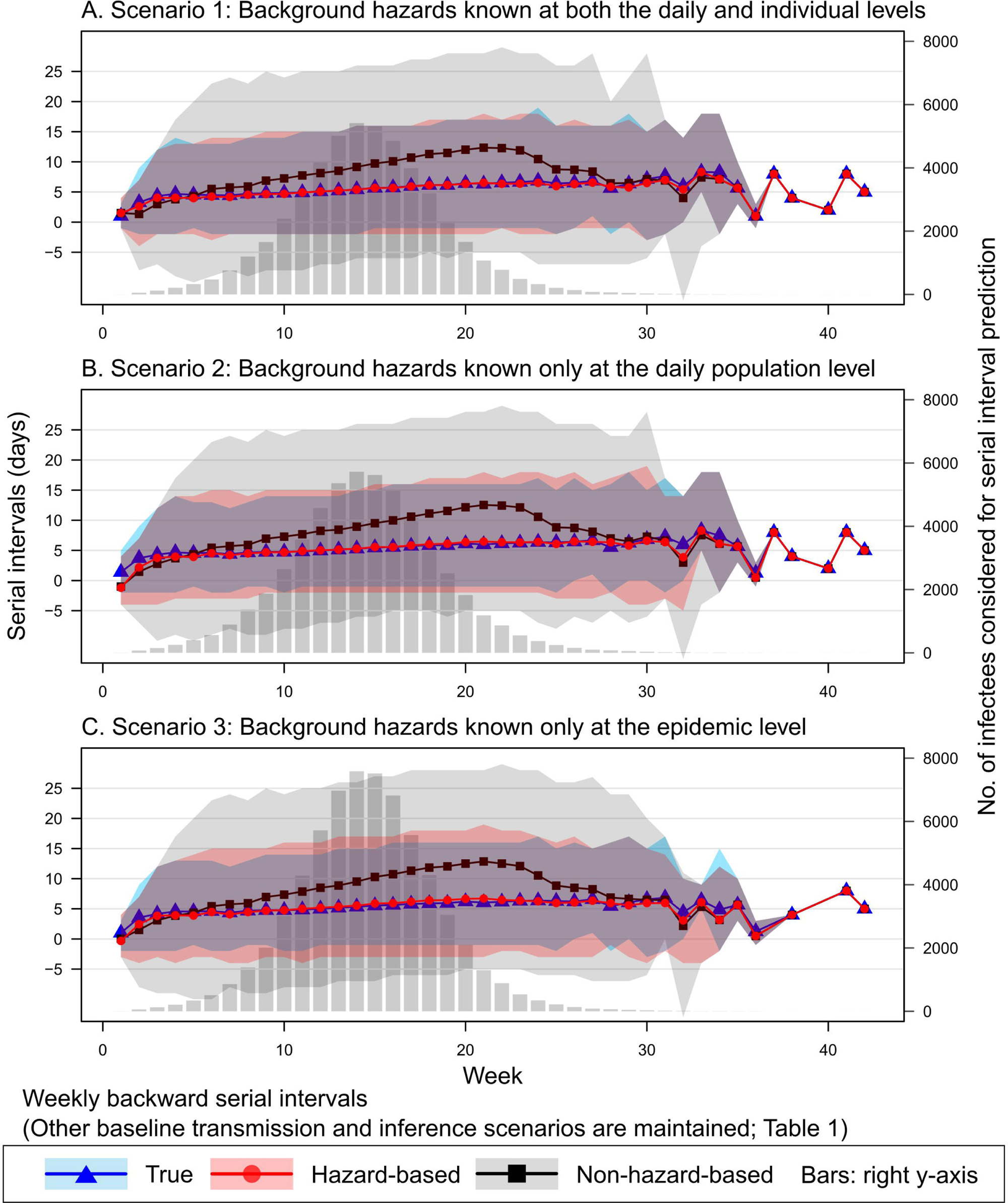
Weekly empirical serial-interval (SI) distributions across background-hazard knowledge scenarios. This figure compares weekly backward SI distributions across three background-hazard knowledge scenarios, and within each scenario, between true, hazard-based, and non-hazard-based reconstruction. The analysis was conducted on exposure-history data generated under the baseline transmission and inference scenarios, with the sole exception being the background-hazard knowledge scenario explored. SIs were computed for infectees who developed symptoms within a given week. Hazard-based SI distributions (red circles and shaded regions) were reconstructed for infectees whose infection was attributed to observed exposures rather than the background hazard. True SI distributions (blue triangles and shaded regions) were reconstructed for the same infectees. Non-hazard-based SI distributions (black squares and shaded regions) were obtained by randomly sampling an observed exposure within an assumed 20-day maximum incubation window for the exact same number of infectees per week. The points and shaded regions represent the median and 95th percentile intervals, respectively. Grey bars, plotted against the secondary right y-axis, indicate the total number of infectees that contributed to the SI reconstruction each week.

**Figure 7.**
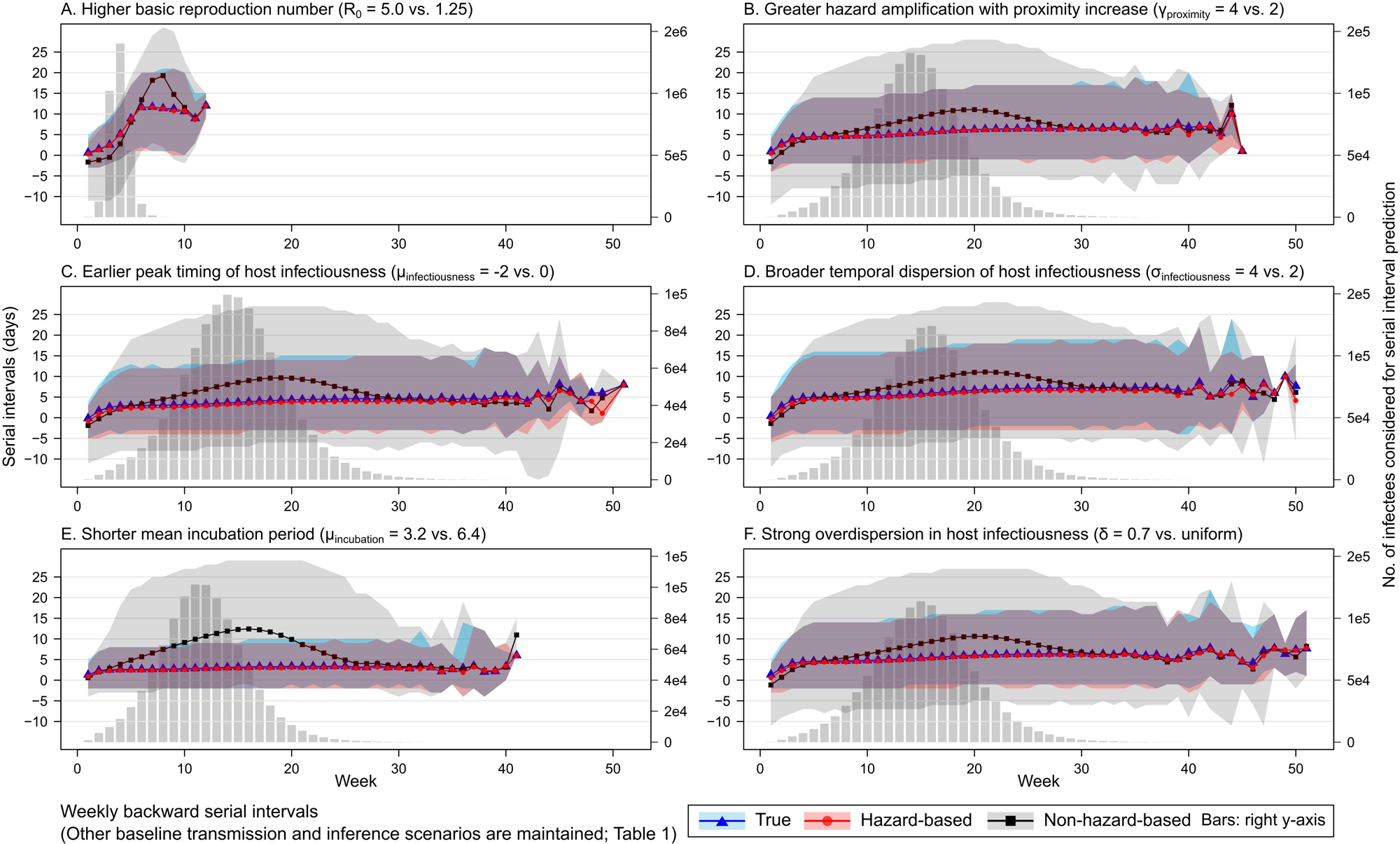
Weekly empirical serial-interval (SI) distributions under alternative transmission scenarios. This figure compares weekly backward SI distributions, evaluating true, hazard-based, and non-hazard-based SI reconstructions across six alternative transmission scenarios. Inference was performed on the exposure-history datasets generated under the baseline transmission and inference scenarios, with the sole exception being the specific transmission scenario explored in each panel. SIs were computed for infectees who developed symptoms within a given week. Hazard-based SI distributions (red circles and shaded regions) were reconstructed for infectees whose infection was attributed to observed exposures rather than the background hazard. True SI distributions (blue triangles and shaded regions) were reconstructed for the same infectees. Non-hazard-based SI distributions were (black squares and shaded regions) obtained by randomly sampling an observed exposure within an assumed 20-day maximum incubation window for the exact same number of infectees per week. Points and shaded regions represent the median and 95th percentile intervals, respectively. Grey bars, plotted against the secondary right y-axis, indicate the total number of infectees that contributed to the SI reconstruction each week.

## 4. Discussion

We demonstrate that parameter estimates from the proposed gradient-based decentralised framework closely match those from centralised exposure-history data, highlighting the viability of decentralised epidemiological inference within future DCT implementations. Gradient-based inference within this framework offers substantially stronger privacy protection than centralised analysis, as the central server receives only securely aggregated gradients, which contain neither exact exposure and infection histories nor direct information capable of revealing transmission chains.

The direction and magnitude of bias in parameter estimates depended on the transmission and inference scenarios. First, when the background hazard was known at the daily population level, bias was close to that in the best-case scenario, in which it was known at both the daily and individual levels. This suggests that daily population-level knowledge of the background hazard, rather than individual-level knowledge, would be sufficient for reliable inference in real-world implementation. Such knowledge could be informed, for example, by daily counts of newly confirmed or symptomatic cases from DCT or independent surveillance systems.

Second, bias decreased as app adoption, case ascertainment, and the completeness of exposure logging increased, highlighting the importance of maximising each and understanding their drivers. Maximising app adoption would require building well-founded public trust through early, direct engagement on privacy-preserving data collection and analysis. Understanding views on centralised and decentralised systems will be important, given survey evidence from the COVID-19 pandemic that demonstrated mixed public attitudes across different nations^24,25^. Such engagement is crucial for balancing the potentially competing interests of safeguarding data privacy and realising the broader public health benefits of analysing these data. Case ascertainment would also benefit from public trust but would remain constrained by disease-specific dynamics (e.g. asymptomatic infections) and operational limitations (e.g. testing capacity). Controlled real-world experiments could quantify how often exposures go unlogged, allowing incomplete logging to be accounted for in inference.

Finally, the bias became larger in transmission scenarios in which infections could be explained by different combinations of parameter values. For example, when most exposures carried a hazard strong enough that transmission was likely regardless of their timing during the infection course, the model lacked the statistical signals needed to identify the peak timing of host infectiousness or the effect of contact proximity. Incorporating prior information on parameters could reduce such bias.

Despite some bias, estimates of the parameters governing the host infectiousness profile remained sufficiently accurate to support downstream inferences, including the SI distribution. The timing of peak host infectiousness relative to symptom onset was inferred to within one day, and bias in the standard deviation was minimal when the background hazard was known at least at the daily population level. This level of accuracy is likely sufficient to inform public health policymaking, such as guidance on the timing and duration of self-isolation. Accurate estimation of the host infectiousness profile enabled reliable hazard-based reconstruction of SI distributions. These reconstructed distributions closely tracked the ground truth, capturing weekly means accurately and matching the 95^th^ percentile intervals reasonably well. This accuracy demonstrates the framework’s broader utility for characterising transmission dynamics, both through direct interpretation of SIs and through their use in estimating reproduction numbers.

It is important to note, however, that the accurate reconstruction of the SI distribution does not imply that individual intervals were always derived from true transmission pairs. Indeed, attribution sensitivity decreased from nearly 90% at 20% app adoption to approximately 50% at full adoption, suggesting that the intervals were increasingly reconstructed from incorrect pairs as observed competing exposures increased. Nevertheless, the accuracy of the reconstructed SI distribution suggests that misattributions were made to epidemiologically comparable exposures, that is, exposures to other infected individuals whose timing relative to symptom onset resembled that of the true infector, thereby preserving the underlying transmission dynamics.

Several factors warrant consideration in future research and implementation. First, our framework will require epidemiological research that informs the form of the hazard function, the background hazard, and the incubation-period distribution among other inputs. Second, our simulation used daily networks in which household contacts occurred once daily and occupational and random contacts more sporadically, simplifying real-world contact patterns. Moreover, contact proximity and duration were sampled without accounting for contact type, whereas in reality household contacts are typically longer and closer than random ones. Third, app adoption and case ascertainment are likely to vary across the population, depending strongly on demographic and socioeconomic factors, leading to clustering of app users and reported cases within particular groups^26–28^. Future studies should therefore incorporate more realistic contact networks and empirical patterns of app usage, case reporting, and other behavioural factors to assess and account for their impact on inference.

Our assessment also did not account for contact settings (e.g. indoor versus outdoor), protective measures (e.g. mask-wearing), or immunity from infection or vaccination. Strategies are needed to record such contextual information, which could impact transmission hazards, and to incorporate it into inference. However, DCT is most useful during early epidemic phases, before substantial population immunity has built up. Thus, while immunity data should still be incorporated, their absence may matter less during these early phases.

Finally, implementing our framework will require in-depth discussion with public health authorities and industry partners, such as Google and Apple, which jointly developed the Google/Apple Exposure Notification (GAEN) system during the COVID-19 pandemic^29^. They will be essential partners for implementation, given the high volume and frequency of sharing of non-identifiable data required between local devices and central servers.

In conclusion, we developed a decentralised framework for epidemiological inference from digital exposure-history data and systematically evaluated its performance across a range of transmission and inference scenarios. Although some scenarios introduced bias into parameter estimates, the framework robustly inferred the host infectiousness profile and the SI distribution, the latter by attributing infections to true infectors or to potential infectors exposed to the case at a similar time relative to their own symptom onset. These findings underscore the potential value of DCT, beyond its primary role as a control measure, for characterising transmission dynamics and informing public health policymaking while preserving user privacy.

## Data Availability

This study used simulated epidemic datasets for epidemiological inference within a decentralised framework. The details of the epidemic simulation are described in the Methods section.

## 5. Acknowledgements

We are grateful to Professor Johannes Abeler (Department of Economics, University of Oxford), Dr Ihsan Kahveci (Leverhulme Centre for Demographic Science, Nuffield Department of Population Health, University of Oxford), Dr Sumali Bajaj (Department of Statistics, University of Oxford), and Alex Yan (Department of Statistics, University of Oxford) for valuable discussions throughout the development of this work.

