## Supplementary Information for "Privacy-Preserving, Decentralised Inference of Epidemic Transmission Dynamics from Digital Exposure Histories"

Younjung Kim^a,b^, Ben Lambert^a,b^, Luca Ferretti^b,c^, Michelle Kendall^b,c,d^, Robert Hinch^b,c^, Jamie Webb^e^, Thomas Rawson^f^, Lucie Abeler-Dörner^b,c^, Peter Horby^b,c^, Michael Parker^e^, Melinda C. Mills^f,g,h^, Christophe Fraser^b,c^, Moritz U.G. Kraemer^b,i^, Christl A. Donnelly^a,b,*^

a Department of Statistics, University of Oxford, Oxford, UK

b Pandemic Sciences Institute, University of Oxford, Oxford, UK

c Nuffield Department of Medicine, University of Oxford, Oxford, UK

d Oxford Martin School, University of Oxford, Oxford, UK

e Ethox Centre, Nuffield Department of Population Health, University of Oxford, Oxford, UK

f Leverhulme Centre for Demographic Science, Nuffield Department of Population Health and Nuffield College, University of Oxford, Oxford, UK

g Department of Economics, Econometrics & Finance, University of Groningen, The Netherlands

h Department of Genetics, University Medical Center Groningen, The Netherlands

i Department of Biology, University of Oxford, Oxford, UK

### 1. Supplementary Methods

#### 1.1. Epidemic Simulation

During the epidemic simulation, we defined a potential infector as an infected individual who poses varying levels of transmission hazard depending on their intrinsic infectiousness over time as well as the proximity and duration of the contacts they make. For a susceptible individual, we defined an exposure as a contact with a potential infector. Building on the formulae used to compute transmission risk scores by the NHS COVID-19 app^1^, the transmission hazard $h_{e}(\tau)$ posed by an exposure $e$ to a potential infector $i$ on day $\tau$ was modelled as:

$$h_{e}(\tau)=exp(\beta_{\text{baseline}}+\beta_{\text{prox}\text{imity}}\cdot\rho_{e})\cdot w(\tau-o_{i})\cdot d_{e}$$

Here, $\rho_{e}$ and $d_{e}$ were the contact proximity and duration of exposure $e$ assigned during network construction. The parameter set $\theta=\{\beta_{\text{baseline}},\beta_{\text{prox}\text{imity}},\mu_{\text{infectiousness}},\sigma_{\text{infectiousness}}\}$, which we aimed to estimate, governed the overall transmissibility of the hypothetical respiratory pathogen (Table 1 in the main text). $\beta_{\text{baseline}}$ represented the log-baseline hazard, and $\beta_{\text{prox}\text{imity}}$ was the coefficient that scaled this hazard based on contact proximity. The term $w(\tau-o_{i})$ represented the intrinsic infectiousness of a potential infector $i$ on day $\tau$, with $o_{i}$ being infector $i$’s symptom onset date. This was modelled using a discretised, peak-normalised Normal distribution $\mathcal{N(}\mu_{\text{infectiousness}},\sigma_{\text{infectiousness}}^{2})$ evaluated over daily intervals, with $\mu_{\text{infectiousness}}$ representing the peak timing of host infectiousness relative to symptom onset date $o_{i}$ and $\sigma_{\text{infectiousness}}$ representing the temporal dispersion of host infectiousness.

The total daily hazard $H_{j}(\tau)$ for any susceptible individual $j$ on day $\tau$ was calculated by aggregating the hazards from all their exposures on that day, denoted by the exposure set $\mathcal{E}_{j}(\tau)$:

$$H_{j}(\tau)=\sum_{e\in\mathcal{E}_{j}(\tau)} h_{e}(\tau)$$

This aggregated hazard directly defined the daily probability of individual $j$ becoming infected $P_{j}(\tau)$ as:

$$P_{j}(\tau)=1-exp(-H_{j}(\tau))$$

To determine whether an infection actually occurred on that day, we performed a Bernoulli trial using $P_{j}(\tau)$. If this trial triggered an infection, the true infector was assigned by sampling from the individual’s exposures $\mathcal{E}_{j}(\tau)$. The probability of attributing the infection to a specific exposure $e$ was equal to the fraction of the total hazard that exposure contributed:

$$\pi_{e}=\frac{h_{e}(\tau)}{H_{j}(\tau)}$$

For each newly infected individual, a symptom onset date was drawn from a discretised Gamma distribution (Table 1). This symptom onset date was then used to determine the intrinsic infectiousness of this individual. Infected individuals were considered no longer infectious after 14 days and subsequently transitioned to a recovered state.

#### 1.2. Epidemiological Inference within a Decentralised Framework

#### 1.2.1. Exposure-History Data Generation by Inference Scenario

To evaluate the impact of varying app adoption levels, we randomly assigned app-user status to individuals and excluded any exposures involving non-app users from the exposure-history data. Assuming a baseline app adoption of 20%, we further investigated the impact of case under-ascertainment and incomplete exposure logging. For case under-ascertainment, a specified proportion of true cases were randomly reclassified as non-cases post-simulation and prior to inference. Consequently, exposures originating from these individuals were excluded from the inference process, despite posing actual transmission risks during the epidemic simulation. For incomplete exposure logging, a set proportion of exposures were randomly deleted from the exposure-history data across all individuals (both cases and non-cases) prior to inference. As a result, these missing exposures were omitted from the inference model, even if they contributed to true transmission risks during the epidemic simulation.

#### 1.2.2. Transmission Hazard Accounting for Unobserved Exposure Hazards

Hazards from unobserved exposures, arising from incomplete app adoption, case ascertainment, and exposure logging constituted the background hazard. During inference, the total daily hazard incorporated the background hazard alongside the hazard from observed exposures as:

$$H_{j}\left( \tau\right)=h_{\text{bg}}\left( \tau\right)+\sum_{e\in\mathcal{E}_{j}^{\text{observed}}(\tau)} h_{e}(\tau)$$

The first term represents the hazard from observed exposures for a susceptible individual $j$ on day $\tau$, defined by the set $\mathcal{E}_{j}^{\text{observed}}$. The second term represents the background hazard from unobserved exposures, a component not included in the hazard function for the epidemic simulation because all hazards were fully specified in the epidemic simulation. To reflect varying levels of information broadcast by the server, we modelled the background hazard under the following three scenarios:

In Scenario 1, individual users’ unobserved hazards are perfectly known:

$$H_{bg,j}(\tau)=\sum_{e\in\mathcal{E}_{j}^{\text{unobserved}}(\tau)} h_{e}(\tau)$$

where $\mathcal{E}_{j}^{\text{unobserved}}(\tau)$ is the set of exposures individual $j$ had with non-app users on day $\tau$. This scenario provides an empirical upper bound for model performance in epidemiological inference.

Scenario 2 set a more realistic situation where daily averaged background hazards are known, but not at the individual level:

$$H_{\text{bg}}(\tau)=\frac{1}{m}\sum_{j=1}^{m} \sum_{e\in\mathcal{E}_{j}^{\text{unobserved}}(\tau)} h_{e}(\tau)$$

where $m$ is the number of app users included in the analysis. During optimisation, $H_{\text{bg}}(\tau)$ was estimated from the same mini-batch $C$ of $m$ users used to compute the likelihood gradients. This approach allows the background hazard to change in response to the epidemic scale, thereby capturing days with higher or lower overall transmission risk. We set Scenario 2 as our baseline scenario when the impact of additional assumptions is assessed. In real-world situations, this type of data could be approximated by other population-level epidemic data, such as prevalence and incidence, although model fitting would require an additional scaling parameter in this case.

Scenario 3 sets the worst-case situation where a flat, uniform background hazard is assumed throughout the study period, ignoring changes between individuals and over days:

$$H_{\text{bg}}(\tau)=c=\frac{1}{m\cdot T}\sum_{j=1}^{m} \sum_{t=1}^{T} \sum_{e\in\mathcal{E}_{j}^{\text{unobserved}}(t)} h_{e}(t)$$

Here, hazards from unobserved exposures are averaged over all $m$ users and all days of the epidemic up to $T$. In real-world situations, this could be approximated by the combined overall level of app non-adoption, case under-ascertainment, and incomplete exposure logging.

#### 1.2.3. Stochastic Variational Inference within a Decentralised Framework

With the above specification of the background hazard, for any susceptible individual $j$, the hazards from both observed and unobserved exposures on day $\tau$ were aggregated to represent the total daily hazard $H_{j}(\tau)$. This, in turn, defined the probability that individual $j$ remains uninfected up to day $t$, expressed as:

$$S_{j}(t)=exp\left( -\sum_{\tau=1}^{t} H_{j}(\tau) \right)$$

Based on the daily survival function $S_{j}(t)$, the marginal probability that individual $j$ becomes infected exactly on day $\tau$ is given by:

$$P_{\text{inf},j}(\tau)=S_{j}(\tau-1)-S_{j}(\tau)$$

where $S_{j}(0)=1$. The specific formulation of the log-likelihood $\log p(\mathcal{D}_{j}|\theta)$ depends on whether individual $j$ is observed as a case ($\mathcal{D}_{j}=\text{case}$) or a non-case ($\mathcal{D}_{j}=\text{non-case}$) at the time of analysis, $T$. First, a case is defined as an app user who is infected and develops symptoms on day $o_{j}\leq T$. (Under the baseline inference scenario, all symptomatic users were identified as cases through the app. See alternative scenarios about case under-ascertainment.) The log-likelihood for a case is constructed as the convolution of the daily infection probability and the probability that the pathogen incubates for exactly $o_{j}-\tau$ days:

$$\log p(\mathcal{D}_{j}=\text{case}|\theta)=log\left( \sum_{\tau=1}^{o_{j}} P_{\text{inf},j}(\tau)\cdot P_{\text{inc}}(o_{j}-\tau) \right)$$

where $P_{\text{inc}}(d)$ is the probability mass function of the discretised Gamma-distributed incubation period evaluated at $d$ days.

Second, a non-case is defined as an app user who does not show symptoms by $T$ for one of two mutually exclusive possibilities: the user (1) has not (yet) been infected or (2) is infected but has not yet developed symptoms. Therefore, the log-likelihood for a non-case is given by:

$$\log p(\mathcal{D}_{j}=\text{non-case}|\theta)=log\left( S_{j}(T)+\sum_{\tau=1}^{T} P_{\text{inf},j}(\tau)\cdot S_{\text{inc}}(T-\tau) \right)$$

where $S_{j}(T)$ represents the probability of remaining uninfected by $T$, and $S_{\text{inc}}(d)$ is the probability that the incubation period exceeds $d$ days. We assumed that the incubation-period distribution was known and provided by the central server, considering that it can be estimated relatively easily from epidemiological survey data.

For these log-likelihoods, local apps must compute the gradients to send back to the server to perform decentralised inference. By the chain rule, the overall gradient is:

$\nabla_{\theta}\log p(\mathcal{D}_{j}|\theta)=\frac{1}{p(\mathcal{D}_{j}|\theta)}\nabla_{\theta}p(\mathcal{D}_{j}|\theta)$.

This, in turn, requires the derivative of the daily infection probability:

$$\nabla_{\theta}P_{\text{inf},j}(\tau)=\nabla_{\theta}S_{j}(\tau-1)-\nabla_{\theta}S_{j}(\tau)$$

where $\nabla_{\theta}S_{j}(t)=-S_{j}(t)\sum_{\tau=1}^{t} \nabla_{\theta}H_{j}(\tau)$

Consequently, the gradient of the log-likelihood inherently depends on the gradient of the daily total hazard $\nabla_{\theta}H_{j}(\tau)$, which is determined by the partial derivatives of the specific observed exposure hazards $h_{e}(\tau)$:

$$\frac{\partial h_{e}(\tau)}{\partial\beta_{baseline}}\&=h_{e}(\tau)$$

$$\frac{\partial h_{e}(\tau)}{\partial\beta_{proximity}}\&=h_{e}(\tau)\cdot\rho_{e}$$

$$\frac{\partial h_{e}(\tau)}{\partial\mu}\&=exp(\beta_{baseline})\cdot d_{e}\cdot exp(\beta_{\text{proximity}}\cdot\rho_{e})\cdot\frac{\partial w(\tau-o_{i})}{\partial\mu}$$

$$\frac{\partial h_{e}(\tau)}{\partial log\sigma}\&=exp(\beta_{baseline})\cdot d_{e}\cdot exp(\beta_{\text{proximity}}\cdot\rho_{e})\cdot\frac{\partial w(\tau-o_{i})}{\partial log\sigma}$$

In a real-world implementation, local apps would transmit only the full gradient vectors $\nabla_{\theta}\log p(\mathcal{D}_{j}|\theta)$ back to the central server. The central server would then aggregate them and use them to update parameters, ensuring that any inference is made without sharing exposure-history data.

We emulated this process by utilising the stochastic variational inference (SVI) approach^2^ paired with the Adam optimiser^3^. Standard Markov chain Monte Carlo (MCMC) approaches are computationally intractable with the decentralised data structure due to their requirement for full-data likelihood evaluations.

Our SVI framework relied on finding a tractable variational distribution $q_{\phi}(\theta)$ that maximises the evidence lower bound (ELBO). To estimate the mean and variance of each global parameter (variational parameters $\phi$) while capturing parameter correlations, we defined $q_{\phi}(\theta)$ as a full-covariance multivariate Normal distribution: $q_{\phi}(\theta\mathcal{)=N(}\mu_{\phi},\Sigma_{\phi})$. We parameterised the covariance matrix via its lower-triangular Cholesky factor $L$, such that $\Sigma_{\phi}=LL^{T}$. In a real-world federated learning system, the server updates the variational parameters $\phi=\{\mu_{\phi},L\}$.

For a total app user population of size $N$ and a sampled federated mini-batch $C$ of size $m$, the ELBO is defined as:

$$\mathcal{L}_{\text{ELBO}}(\phi)=\mathbb{E}_{q_{\phi}(\theta)}\left[ \frac{N}{m}\sum_{j\in C} \log p(\mathcal{D}_{j}|\theta)+log p(\theta) \right]\mathcal{+H(}q_{\phi})$$

where $p(\theta)$ is the joint prior distribution, $\frac{N}{m}$ scales the mini-batch to act as an unbiased population estimator, and $\mathcal{H(}q_{\phi})$ is the analytical entropy of the multivariate Normal distribution^4^:

$$\mathcal{H(}q_{\phi})=\frac{D}{2}\left( 1+log2\pi\right)+\sum_{d=1}^{D} \log L_{dd}$$

with $D=4$ representing the number of global parameters, and $L_{dd}$ representing the diagonal elements of the Cholesky matrix.

To permit gradient-based optimisation through the stochastic expectation $\mathbb{E}_{q_{\phi}}$, we analytically emulated the iterative server-app communication cycle using the path-wise gradient estimator as follows:

First, acting as the central server, we drew $S=3$ independent standard Normal samples $z^{(s)}\mathcal{\sim N(}0,I)$ and deterministically generated the parameters as $\theta^{(s)}=\mu_{\phi}+Lz^{(s)}$. Mimicking the server broadcast, we passed these sampled parameters to the individual-level data subset.

Next, acting as the local apps, we computed the log-likelihood gradients $\nabla_{\theta}\log p(\mathcal{D}_{j}|\theta^{(s)})$ for the individuals in the current mini-batch. This computation was based on the sampled parameters, the local exposure-history data, and the pre-programmed hazard function, conditioned appropriately on each user’s observed case status.

Finally, acting again as the central server, we aggregated these gradients to estimate the overall gradient of the ELBO with respect to the variational parameters $\phi$:

$$\nabla_{\phi}\mathcal{L}_{\text{ELBO}}\approx\frac{1}{S}\sum_{s=1}^{S} \nabla_{\phi}\theta^{(s)}\cdot\left[ \frac{N}{m}\sum_{j\in C} \nabla_{\theta}\log p(\mathcal{D}_{j}|\theta^{(s)})+\nabla_{\theta}\log p(\theta^{(s)}) \right]+\nabla_{\phi}\mathcal{H(}q_{\phi})$$

Based on these gradients, we updated the variational parameters $\phi$ using the Adam optimiser. In our emulation, this continuous server-app communication cycle was iterated for 5,000 steps, and the final parameter estimates were obtained by averaging over the final 10% of the optimisation trajectory. Visual inspection indicated that the variational means had stabilised well before the averaging window and across all simulated datasets.

#### 1.2.4. Secure Aggregation of Gradient Values

For a mini-batch of $n$ users, random masks drawn from a wide uniform distribution were assigned to $m-1$ users, and the $m$-th user was assigned the negative sum of the preceding masks. This zero-sum masking ensures that the masks perfectly cancel out across the mini-batch, yielding aggregate masked gradients that are identical to the aggregate raw gradients, thereby producing the same inference results. In practice, masks are generated pairwise between users through cryptographic key agreement to prevent no single party, including a central server, from identifying any individual user’s mask^5^.

| **Table S1.** Toy example of secure aggregation | | | |
| --- | --- | --- | --- |
|  | Raw gradient (kept on device) | Mask^a^ | Masked gradient (sent to server) |
| User 1 (case) | + 0.8 | − 41.6 | − 40.8 |
| User 2 (non-case) | − 0.3 | + 63.1 | + 62.8 |
| User 3 (non-case) | − 0.2 | − 21.5 | − 21.7 |
| Sum | + 0.3 | 0.0 | + 0.3 |
| a The masks for users 1 and 2 are drawn from U(−100, 100) and the mask for user 3 is set so that all three sum to zero. | | | |

#### 1.2.5. Benchmarking Decentralised Inference against Standard MCMC

To assess the accuracy and reliability of our decentralised inference approach, we conducted a benchmarking analysis against a Hamiltonian Monte Carlo (HMC) method. Unlike our federated approach, standard HMC works by evaluating the exact, full-data likelihood, which necessitates the centralisation of exposure-history data^6^.

HMC was performed using the rstan package^7^, employing an equivalent cohort sample size and identical weakly informative prior distributions to those used in the SVI approach: Normal distributions with mean 0 and standard deviation 5 for $\beta_{\text{baseline}},\beta_{\text{prox}\text{imity}}$, and $\mu_{\text{infectiousness}}$, and with mean 0 and standard deviation 2 for log $\sigma_{\text{infectiousness}}$. For each simulated dataset, we ran two chains of 1000 iterations each, including 500 warm-up iterations, and assessed convergence based on visual inspection of the trace plots, the Gelman-Rubin diagnostic (<1.01), and the effective sample size (>400).

We evaluated both approaches with the baseline parameter values at the lowest explored bound of app adoption (20%). This lower bound was selected as a rigorous stress test; algorithmic discrepancies between variational approximations and exact MCMC sampling can be most pronounced under conditions of high data sparsity and uncertainty^8,9^. As our performance metric, we computed the difference between the SVI and HMC estimates for both the posterior means and standard deviations across all 100 simulations.

### 2. Supplementary Figures

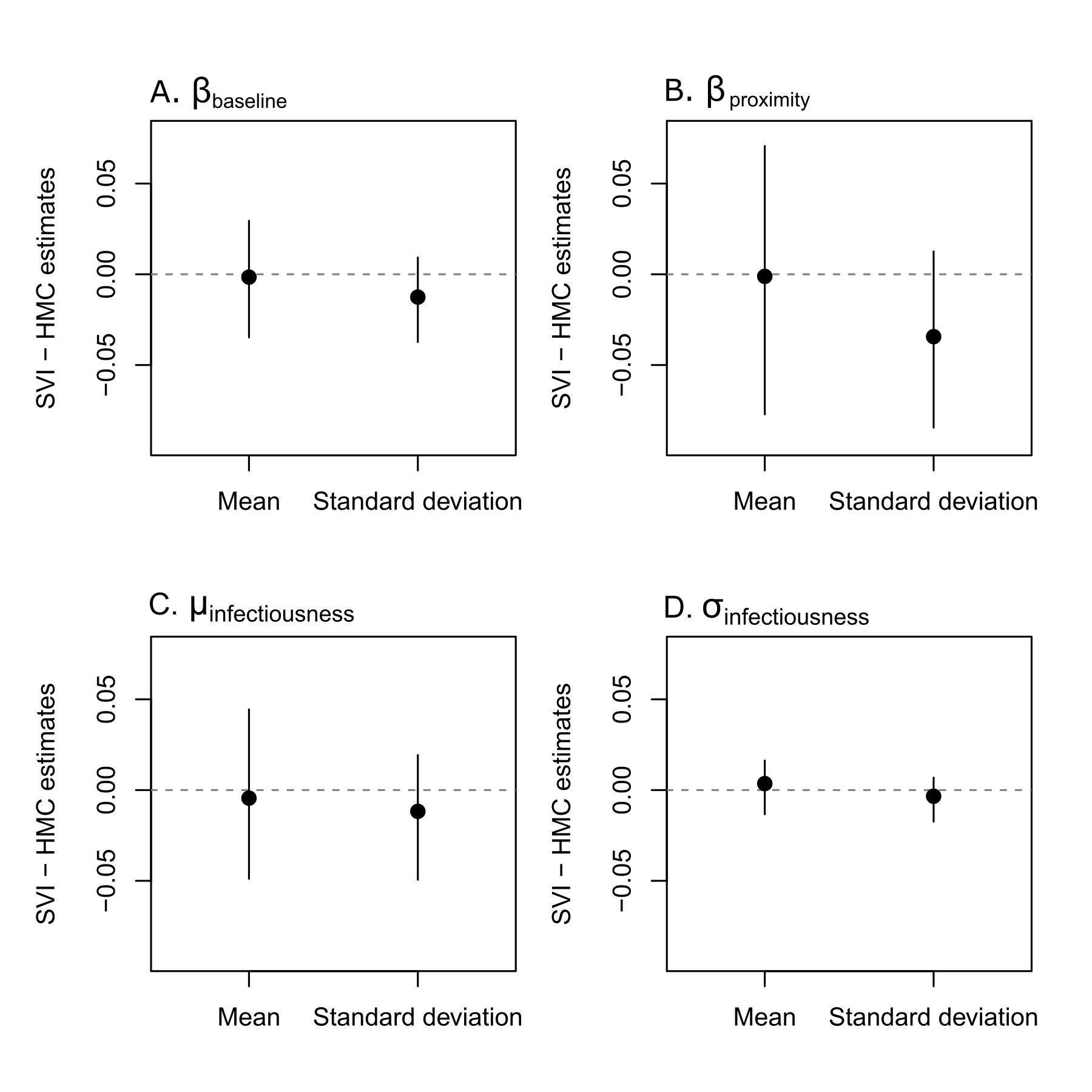

**Figure S1. Comparison of parameter estimates derived from stochastic variational inference (SVI) versus Hamiltonian Monte Carlo (HMC).** SVI estimates were obtained within a decentralised framework using log-likelihood gradients, whereas HMC estimates were obtained by fitting a model directly to centralised exposure-history data using rstan. This figure shows the performance of SVI against an HMC method across four key parameters: (A) $\beta_{baseline}$, the log baseline hazard per day of contact; (B) $\beta_{proximity}$, the log hazard ratio per unit increase in the proximity score; (C) $\mu_{infectiousness}$, the timing of peak host infectiousness relative to symptom onset (days); and (D) $\sigma_{infectiousness}$, the temporal dispersion of host infectiousness (days). The inference was made under the baseline transmission and inference scenario, assuming 20% adoption and that hazards from unobserved exposures were known at the daily population level (background-hazard scenario 2). For each simulated dataset, the difference between the SVI and HMC estimates was calculated for both the mean and the standard deviation. Black points denote the mean differences across all simulations, while the vertical lines indicate the 95^th^ percentile intervals.

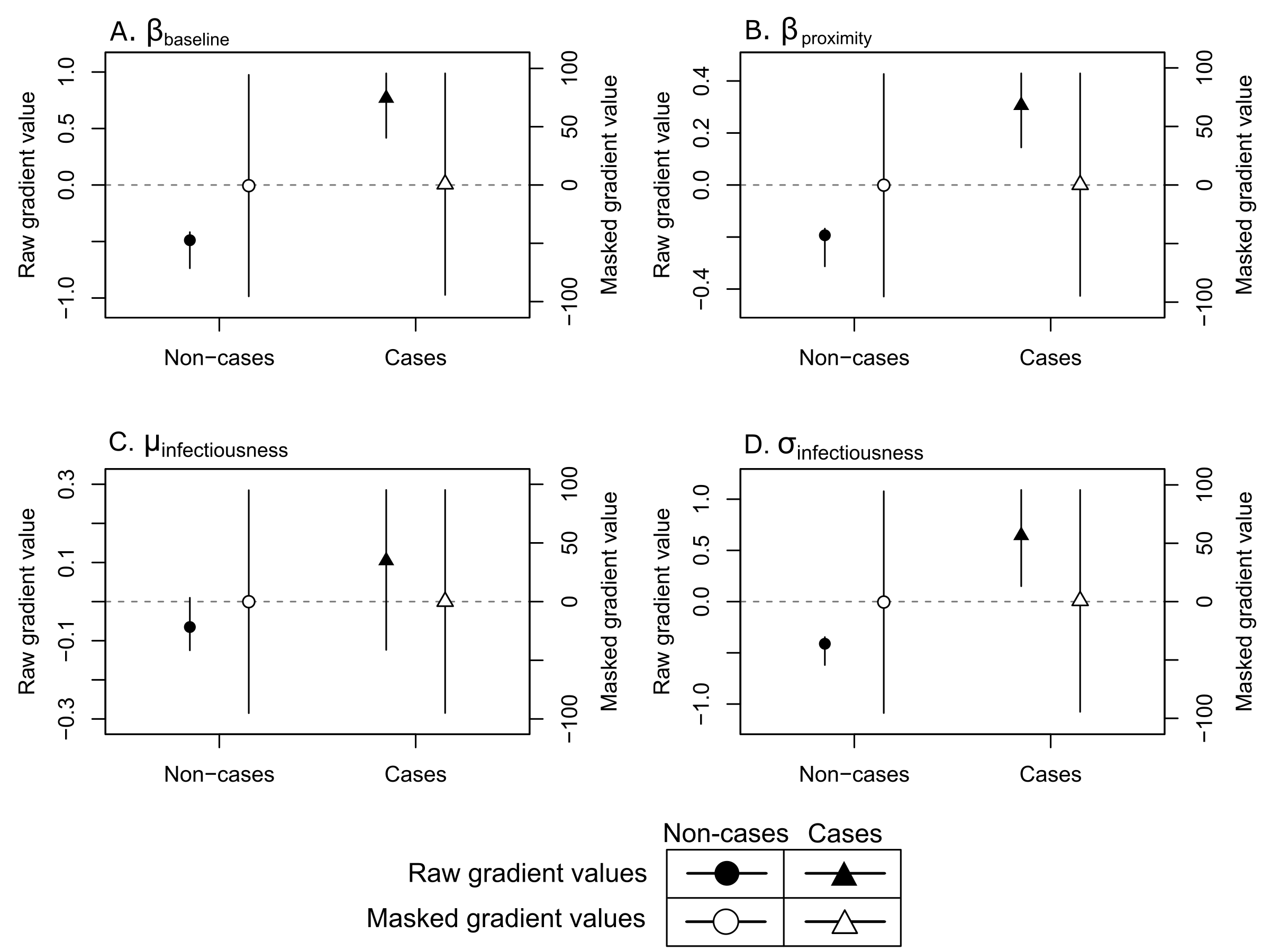

**Figure S2. Privacy preservation of individual gradient values via secure aggregation.** The panels display the median and 95^th^ percentile intervals of individual gradient values at the final stochastic variational inference (SVI) iteration, pooled across 100 simulations under the baseline transmission and inference scenarios, for four transmission parameters: (A) $\beta_{baseline}$, the log baseline hazard per day of contact; (B) $\beta_{proximity}$, the log hazard ratio per unit increase in the proximity score; (C) $\mu_{infectiousness}$, the timing of peak host infectiousness relative to symptom onset (days); and (D) $\sigma_{infectiousness}$, the temporal dispersion of host infectiousness (days). Solid symbols (circles for non-cases, triangles for cases) represent raw, unmasked gradients corresponding to the left y-axis. Open symbols represent masked gradients derived by emulating a secure aggregation process within a decentralised framework, corresponding to the right y-axis.

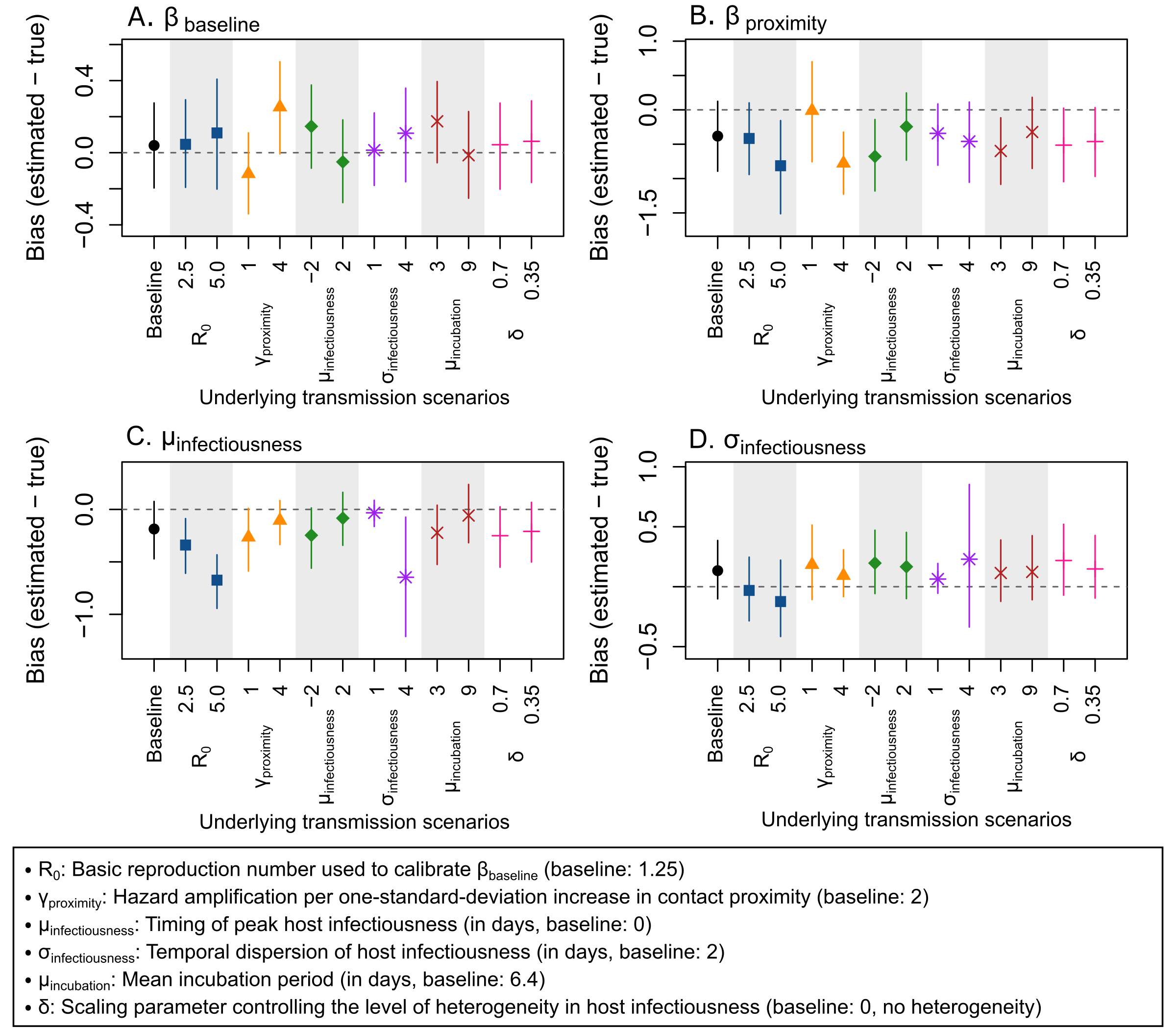

**Figure S3. Impact of alternative transmission scenarios on parameter estimation.** The x-axis represents the alternative transmission scenarios used to simulate the exposure-history datasets, while the y-axis shows the resulting bias, calculated as the difference between the estimated mean and the true parameter values. Points and vertical lines represent the median and 95^th^ percentile intervals of this bias across 100 simulated exposure-history datasets. Inference was conducted under the baseline transmission and inference scenarios, with the only exception being the specific alternative transmission scenario evaluated on the x-axis.

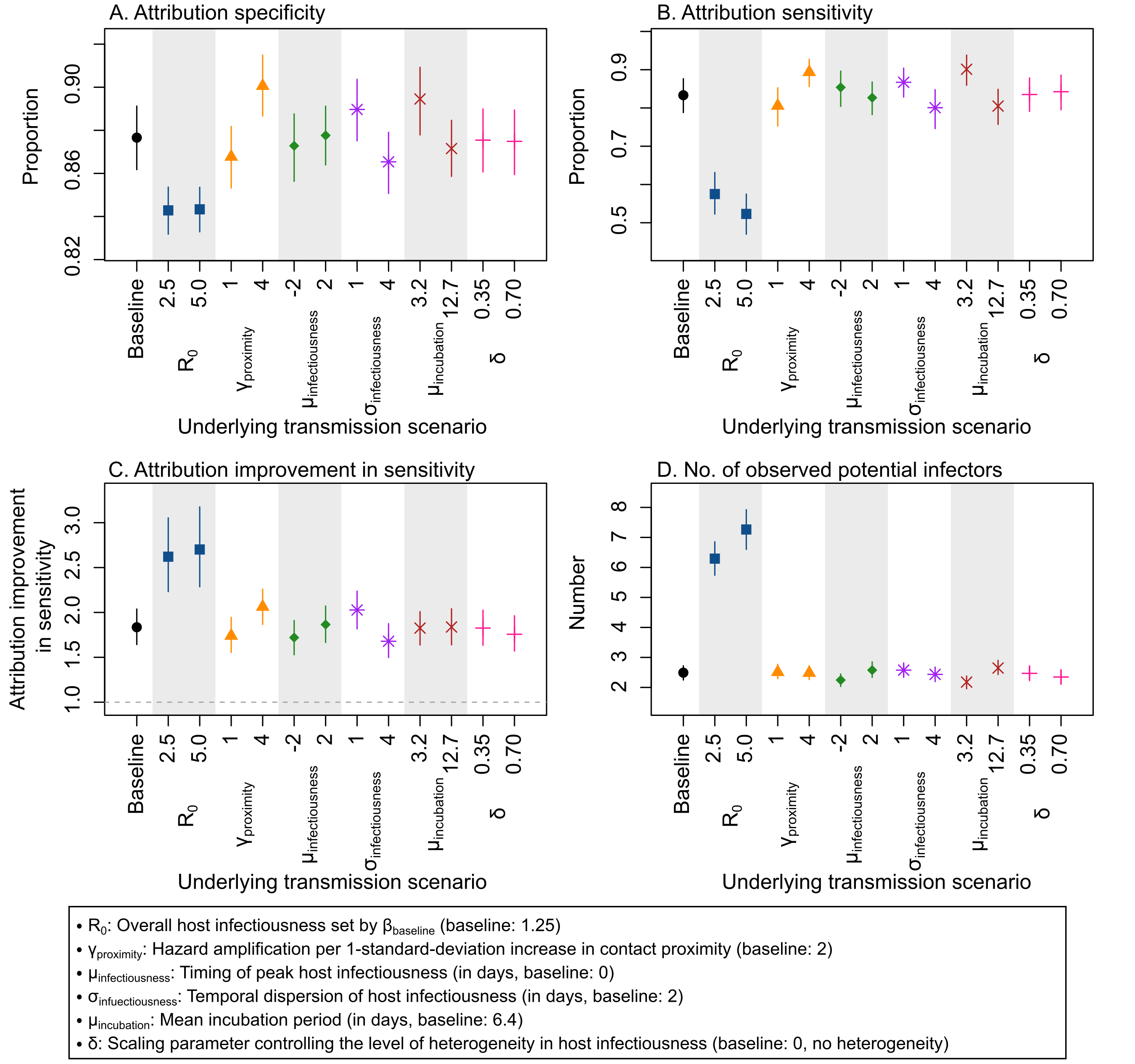

**Figure S4. Impact of alternative transmission scenarios on infector attribution.** This figure shows the performance of infector attribution across alternative transmission scenarios: (A) attribution specificity (attribution to the background hazard when the true infector is unobserved); (B) attribution sensitivity (attribution to the true infector when attribution is made to observed exposures); (C) attribution improvement in sensitivity (improvement in sensitivity for hazard-based attribution relative to non-hazard-based attribution); and (D) the number of observed potential infectors. The x-axis represents the specific transmission scenarios used to simulate the exposure-history datasets. Points and vertical lines represent the median and 95^th^ percentile intervals of these metrics across 100 simulated exposure-history datasets. Inference was conducted under the baseline transmission and inference scenarios, with the only exception being the specific alternative transmission scenario evaluated on the x-axis.

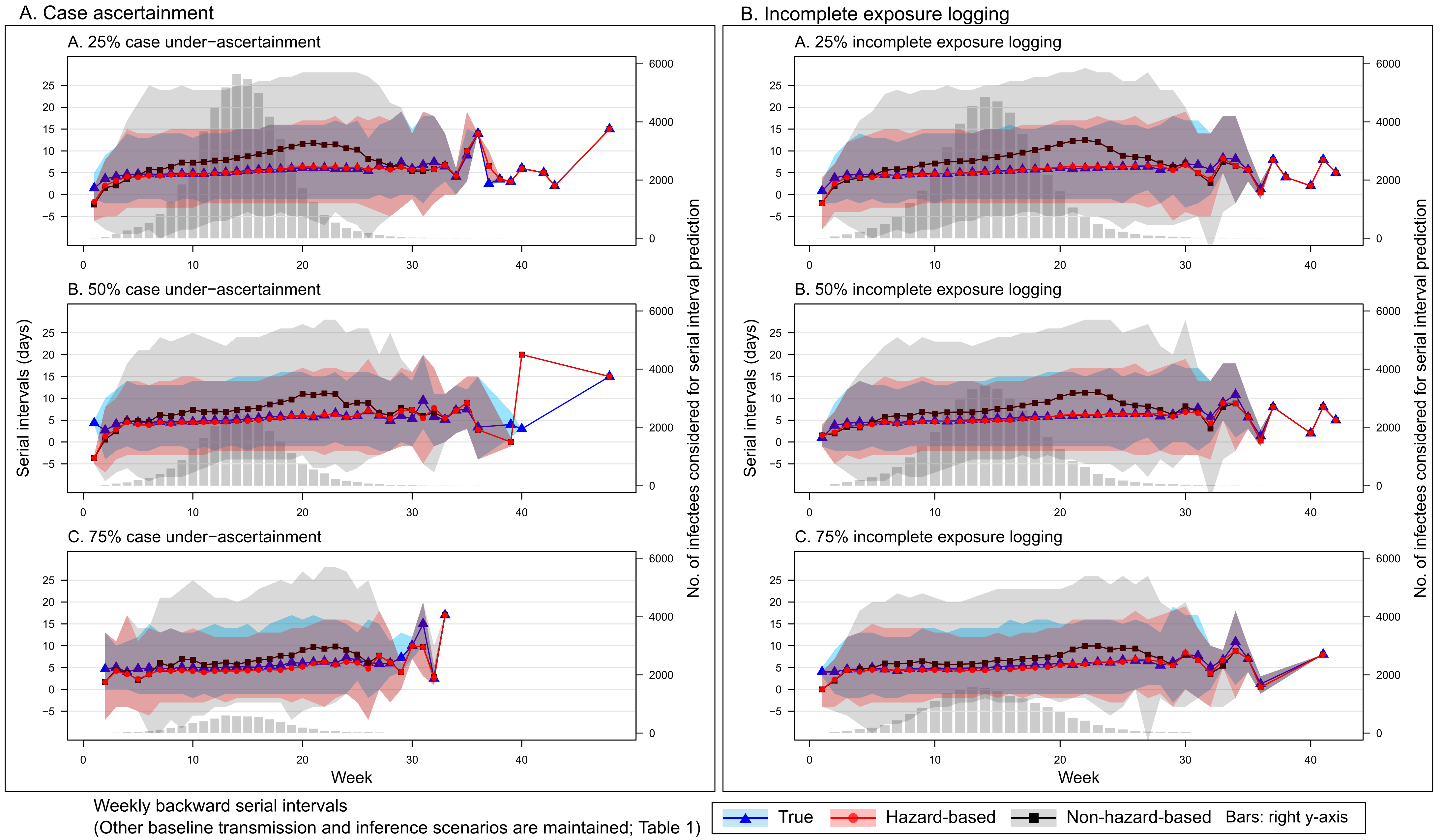

**Figure S5. Impact of case under-ascertainment and incomplete exposure logging on the reconstruction of serial-interval (SI) distributions.** This figure compares weekly backward SI distributions, evaluating true, hazard-based, and non-hazard-based SI reconstructions across different levels of case under-ascertainment or incomplete exposure logging. At a given level of case under-ascertainment or incomplete exposure logging, the inference was conducted under the baseline transmission and inference scenarios, assuming a 20% app adoption and that hazards from unobserved exposures were known at the daily population level (background-hazard scenario 2). SIs were computed for infectees who developed symptoms within a given week. Hazard-based SI distributions (red circles and shaded regions) were reconstructed for infectees whose infection was attributed to observed exposures rather than the background hazard. True SI distributions (blue triangles and shaded regions) were reconstructed for the same infectees. Non-hazard-based SI distributions were (black squares and shaded regions) obtained by randomly sampling an observed exposure within an assumed 20-day maximum incubation window for the exact same number of infectees per week. Points and shaded regions represent the median and 95th percentile intervals, respectively. Grey bars, plotted against the secondary right y-axis, indicate the total number of infectees that contributed to the SI reconstruction each week.
